# Monkeypox self-diagnosis abilities, determinants of vaccination intention and self-isolation intention after diagnosis among MSM in the Netherlands

**DOI:** 10.1101/2022.07.29.22278167

**Authors:** Haoyi Wang, Kennedy J.I. d’Abreu de Paulo, Thomas Gültzow, Hanne M.L. Zimmermann, Kai. J. Jonas

**Affiliations:** Department of Work and Social Psychology, Maastricht University, Maastricht, The Netherlands

## Abstract

Monkeypox is a zoonotic disease and leads to a smallpox-like disease in humans. The current epidemic in European countries requires informed responses. We investigated the ability to self-diagnose a potential monkeypox infection, and determinants of vaccination intention and self-isolation intention after exposure among MSM in the Netherlands. We found that about half were able to self-diagnose monkeypox, that 72% had a high intention to get vaccinated and 44% to self-isolate after monkeypox exposure. Determinants went beyond mere risk behaviour criteria.

**Data availability:** Data is available upon request.

**Ethical statement:** The study was assessed and approved by the ERCPN of Maastricht University (ref.188_11_02_2018_S32). Informed consent was provided by all participants.

**Funding:** There was no funding source for this study.

**Authors’ contributions:** All authors conceptualised this research; KJJ collected the data for this research; HW and KJIDDP analysed the data; all authors drafted the manuscript; all authors critically revised the manuscript for intellectual content; All authors read and approved the final version of the manuscript.

## Introduction

Monkeypox is a zoonotic disease which is caused by an orthopoxvirus and leads to a smallpox-like disease in humans [1]. The global epidemic has recently changed from infections predominantly due to an interaction or activity with animals to human-to-human transmission [1, 2]. The monkeypox epidemic in European countries is currently predominantly affecting men-who-have-sex-with-men (MSM) and number of infections are still increasing [3, 4]. In the Netherlands currently 818 cases (25.07.2022) have been reported, with the majority in Amsterdam.

Given the incubation period of 9 days in the case of invasive exposure [5], it is important to detect infections quickly, which rests the responsibility to detect the typical lesions on affected individuals. However, self-diagnosis of novel diseases can be challenging. Furthermore, infected individuals should isolate for up to 21 days until all symptoms are gone [4]. Especially after all COVID-19 related measures, there is limited data on the acceptance of such measures. At the same time, vaccination programs have commenced in many countries that focus on high-risk populations, for example MSM who are using HIV pre-exposure prophylaxis (PrEP) [6]. Given vaccine scarcity [7], it is paramount to identify highest at-risk populations, but also to gauge general vaccination intention in moderate at-risk populations.

To our knowledge neither findings on the ability of at-risk individuals to diagnose a potential monkeypox infection themselves and seek medical help, nor determinants of vaccination intention and self-isolation intention after diagnosis among MSM in the Netherlands have been reported. Yet, these results are relevant to inform further national responses above and beyond current focal populations. Therefore, we aimed to investigate the ability to self-diagnose monkeypox skin lesions, the intention to get vaccinated (hereafter vaccination intention) and the intention to self-isolate after a monkeypox infection (hereafter self-isolation intention).

## Monkeypox self-diagnosis, vaccination intention, and self-isolation intention after diagnosis

We conducted an online survey among MSM using a cohort established in 2017 (n=257), along with recruitment of MSM on a gay online dating app (n=137) in the first half of July 2022, prior to the start of structural monkeypox vaccination in the Netherlands [8]. Of the included 394 MSM, 43% were below the age of 45-years, 6% were living with HIV and 66% were currently using PrEP (see supplementary materials S1 for the population characteristics by PrEP use status).

We provided participants with four images and asked them to indicate what condition this could be. All of the images were showing lesions in parts of the face, one depicted a monkeypox lesion, the other three a staphylococcus infection, a syphilis stage-2, and eczema. Only the image of eczema was diagnosed predominantly correct, monkeypox and staphylococcus images triggered correct hits, but also considerable amounts of false positives, syphilis stage-2 was most frequently misdiagnosed as monkeypox (Table 1).

**Table 1.** Ability to self-diagnose a monkeypox lesion.

| Shown/diagnosed as | Monkeypox (%) | Staphylococcus (%) | Syphilis (%) | Eczema (%) |
| --- | --- | --- | --- | --- |
| Monkeypox | 52.3 | 38.1 | 7.6 | 2.0 |
| Staphylococcus | 23.9 | 42.4 | 26.6 | 7.1 |
| Syphilis stage-2 | 51.8 | 23.1 | 23.1 | 2.0 |
| Eczema | 3.6 | 24.9 | 5.1 | 66.5 |

Overall, 70% showed high vaccination intention and 44% showed high self-isolation intention. Given that current monkeypox vaccinations are administered to PrEP-using MSM in the Netherlands [6], we adjusted for PrEP use status (current users (n=260) vs. PrEP-naïve MSM or PrEP-discontinued MSM(n=140)) to compare the standardized prevalence ratio (SPR). We found that despite of the higher prevalence among PrEP users in both vaccination intention and self-isolation intention, the adjusted SPRs showed no significant differences, indicating similar vaccination and self-isolation intentions among PrEP users and non-PrEP-users (Table 2).

**Table 2.** Prevalence and standardized prevalence ratio of Monkeypox vaccination intention and self-isolation intention.

| Sub-population | vaccination intention<br>(Extremely likely vs. rest of scale) |  |  |  | monkeypox self-isolation intention<br>(Extremely likely vs. rest of scale) |  |  |  |
| --- | --- | --- | --- | --- | --- | --- | --- | --- |
|  | Prevalence (%) | 95%CI | SPR | 95%CI | Prevalence (%) | 95%CI | SPR | 95%CI |
| PrEP users | 73.86 | 67.97;79.00 | 1.01 | 0.87;1.17 | 43.57 | 37.45;49.81 | 1.06 | 0.87;1.28 |
| Non-PrEP users | 71.31 | 62.73;78.59 | 0.97 | 0.78;1.19 | 35.25 | 27.34;33.06 | 0.86 | 0.63;1.14 |
| Total sample | 70.01 | 65.35;74.36 | NA | NA | 43.91 | 39.09;48.84 | NA | NA |

Note: \* adjusted for PrEP use status. NA = not applicable.

## Determinants predicting vaccination and self-isolation intention

To identify vaccination intention differences and among MSM sub-populations and which sub-population may follow the current self-isolation policy best, we conducted two multivariable logistic regression analyses with socio-demographic, behavioural, and psychosocial determinants. First, we conducted univariable logistic modelling with each of the selected determinants to investigate potential correlations with being extremely likely to get vaccinated. We retained all determinants with p<0.10 in the multivariable model, given the relatively small sample size (see Table 3 for included determinants and models’ details). In a sensitivity analysis, we combined somewhat- and extremely-likely intentions to define high intention to explore the implications on the endpoint selection (see supplementary materials S2).

**Table 3.**
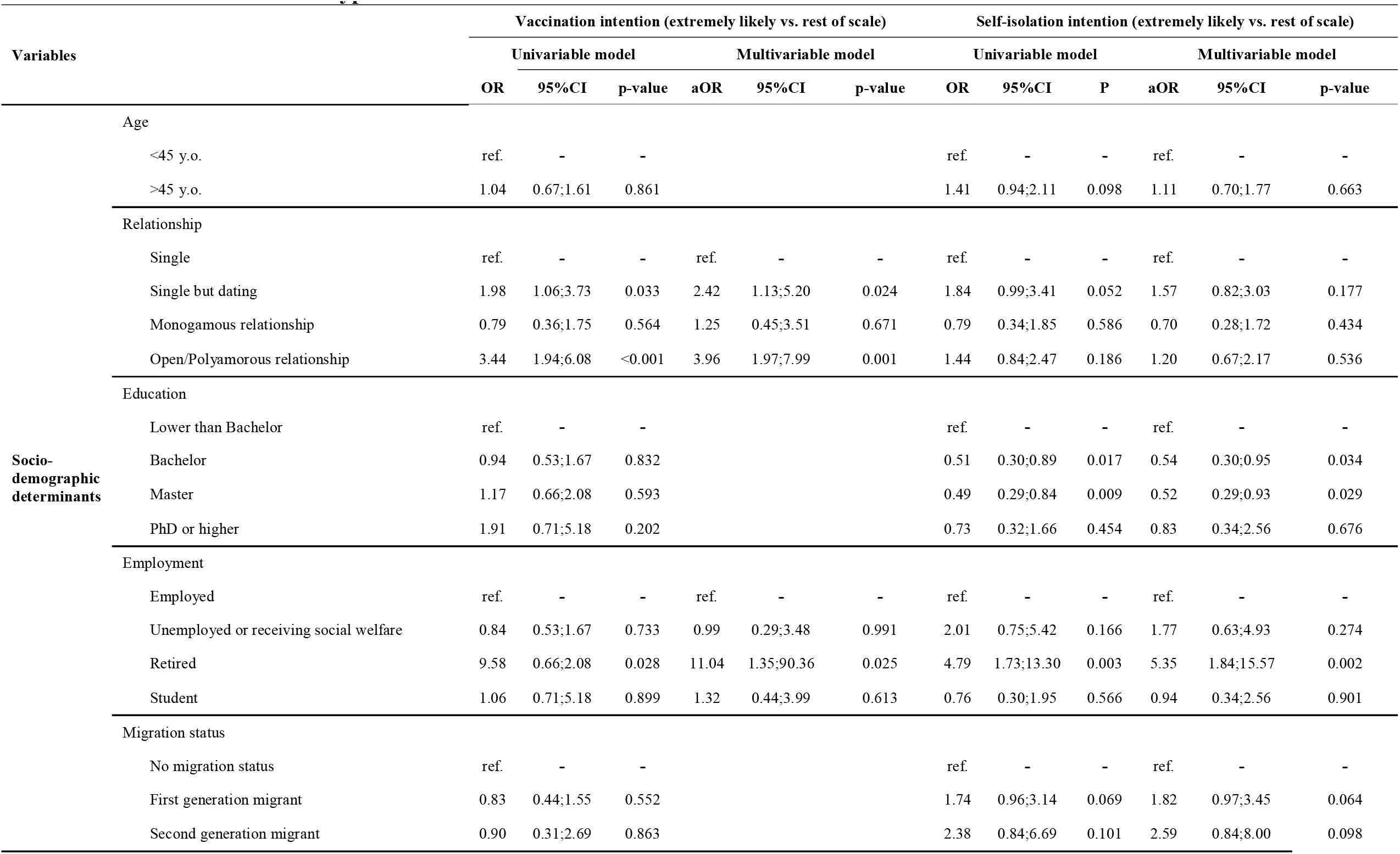
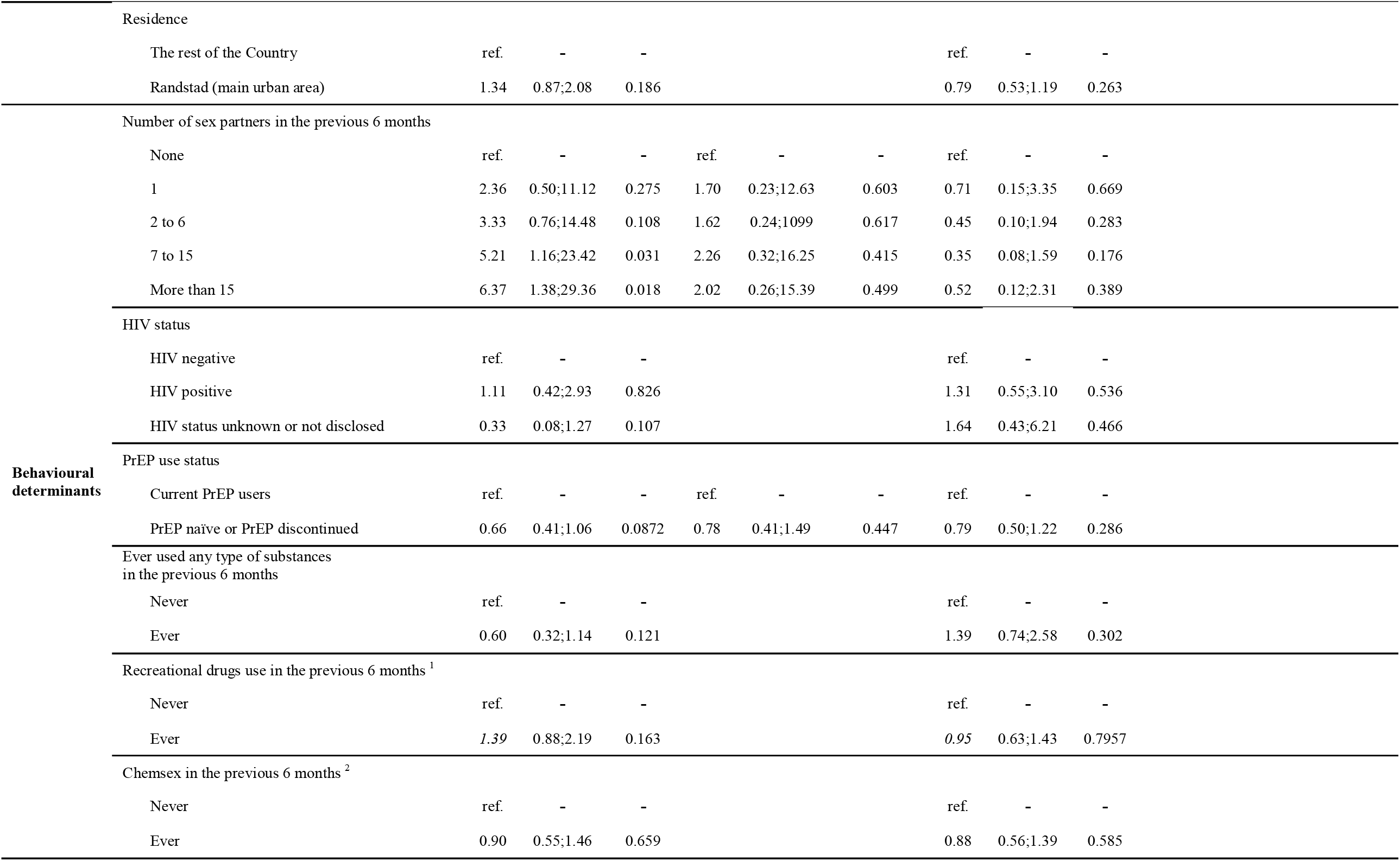
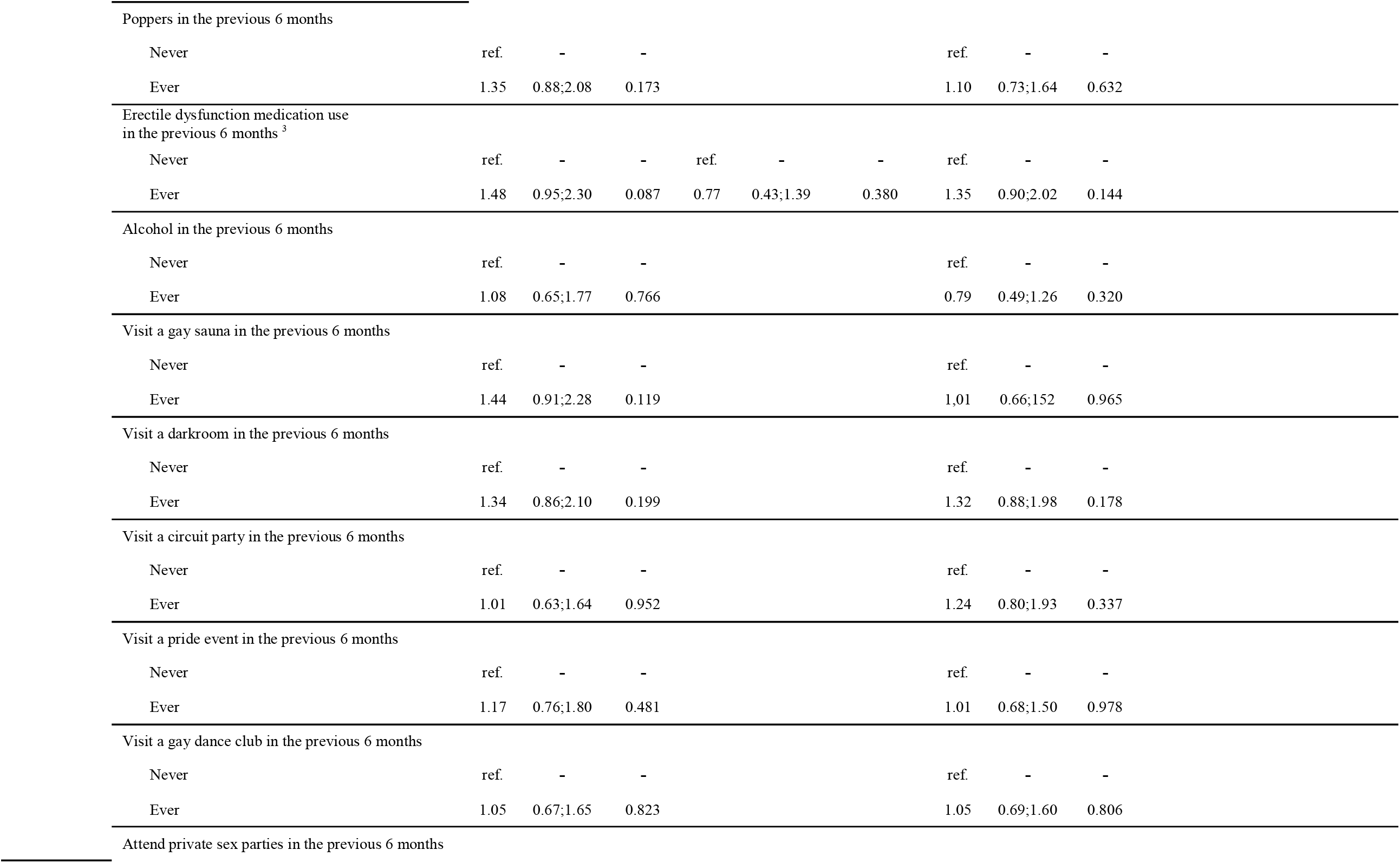
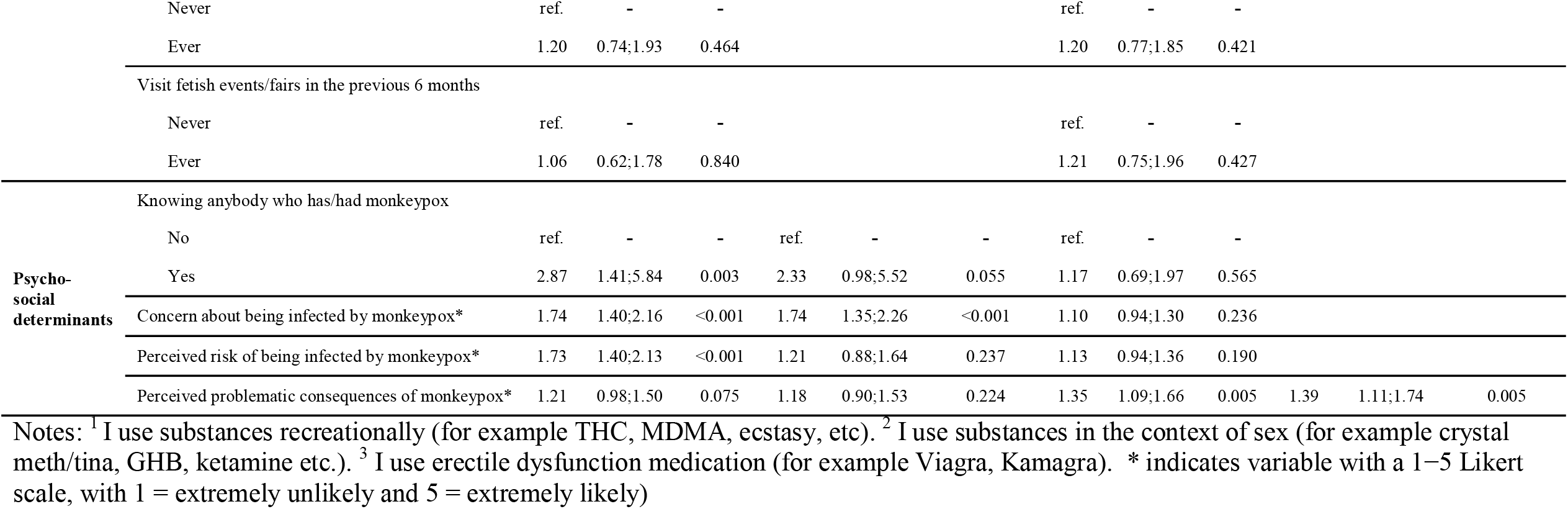
Determinants of Monkeypox vaccination intention and self-isolation intention.

For vaccination intention as endpoint, among socio-demographic determinants, MSM who were single but dating (adjusted odds ratio (aOR)=2.42), who had an open/polyamorous relationship (aOR=3.96) and who were retired (aOR=11.04) were more likely to have high vaccination intentions. No behavioural determinants were found to be statistically associated. Among psycho-social determinants, we found that knowing someone who has/had monkeypox (aOR=2.33) and being worried about a monkeypox infection (aOR=1.74) was associated with high vaccination intention.

For self-isolation intention as endpoint, almost all included socio-demographic determinants showed relevant associations. MSM with bachelor (aOR=0.54) and master (aOR=0.52) degrees were less likely to self-isolate after exposure for 21 days. Those who were retired (aOR=5.35) and who were first-generation (aOR=1.82) and second-generation (aOR=2.59) migrants showed higher intentions on the other hand. Similar to the vaccination intention, no behavioural determinant was found to be associated with high self-isolation intentions. Among psycho-social determinants, MSM who perceived more problematic consequences due to a monkeypox infection (aOR=1.39) were more likely to self-isolate.

## Discussion

The findings of 394 MSM in the Netherlands showed that about half were able to self-diagnose Monkeypox from other skin lesions, and that overall, 70% had a high intention to get vaccinated and 44% to self-isolate after Monkeypox diagnosis. Each of these outcomes hold relevant implications to guide national responses.

First, both the ability to self-diagnose and intention to self-isolate are relatively low. Since public health and social measures are considered essential [9] to reduce transmission of pathogens with epidemic potential, health education campaigns/interventions should not only aim to increase vaccination intentions, but also to support people in following other mitigation regulations such as isolation.

Second, although sexual risk is the most frequently suspected route of infection [10] and is the main reason for the prioritization to be vaccinated in the Netherlands since July 25th [8] high intention to get vaccinated and self-isolate in this study were not predicted by any of the sexual risk behaviours. This contradicts the current public communication and vaccination strategy that focuses on motivating those showing higher sexual risk behaviours, while in fact certain socio-demographic (e.g., education level and migration status) and psycho-social variables (e.g., knowing someone that has/had monkeypox and worrying) were found more predictive. Our results suggest that increasing our efforts to also reach highly educated and non-migrant or Western MSM with these messages might therefore be more important, as well as presenting realistic stories of monkeypox infections using peer-communication without inducing fear (even though our findings imply that it could work [11]).

Also, since vaccination intentions did not differ between PrEP users and non-PrEP-users and non-PrEP-users also show multiple risk behaviours, prioritizing PrEP users may not be sufficient to reach the full potential of the vaccine. Finally, albeit most monkeypox cases have been reported from Amsterdam, our investigation of geographic differences did not yield significant results.

## Conclusions

To conclude, based on our findings governments and public health services should not only focus on vaccinations in their communication but should also aim to increase MSMs’ intentions to isolate and ability to accurately self-diagnose. Efforts should also be stepped up to target MSM at the highest risk, especially those with little concern about monkeypox and those with a high level of education without a (non-Western) migration background.

## Data Availability

All data produced in the present study are available upon reasonable request to the authors.

## Acknowledgements

We thank all study participants contribute to our study.

## Conflict of interest

None declared.

## Notes

### Competing Interest Statement

The authors have declared no competing interest.

### Author Declarations

The study was assessed and approved by the ERCPN of Maastricht University (ref.188_11_02_2018_S32). Informed consent was provided by all participants.

