## Supplementary materials for "Monkeypox self-diagnosis abilities, determinants of vaccination intention and self-isolation intention after diagnosis among MSM in the Netherlands"

**Table S1. Study characteristics by total sample and by PrEP use status.**

| **Variables** | | | **Total sample (n=394)** | | **PrEP (n=241)** | | **Non-PrEP (n=122)** | | **Chi-square test (p-value)** |
| --- | --- | --- | --- | --- | --- | --- | --- | --- | --- |
|  |  |  | **N** | **%** | **N** | **%** | **N** | **%** |  |
| Socio-demographic determinants | Age |  |  |  |  |  |  |  |  |
|  |  | <45 y.o. | 171 | 43.4 | 90 | 37.34 | 69 | 56.56 | <.001 |
|  |  | >45 y.o. | 223 | 56.6 | 151 | 62.66 | 53 | 43.44 |  |
|  | Relationship |  |  |  |  |  |  |  |  |
|  |  | Single | 79 | 20.05 | 47 | 19.5 | 27 | 22.13 | <.001 |
|  |  | Single but dating | 91 | 23.1 | 58 | 24.07 | 26 | 21.31 |  |
|  |  | Monogamous relationship | 35 | 8.88 | 6 | 2.49 | 27 | 22.13 |  |
|  |  | Open/Polyamorous relationship | 189 | 47.97 | 130 | 53.94 | 42 | 34.43 |  |
|  | Education |  |  |  |  |  |  |  |  |
|  |  | Lower than Bachelor | 89 | 22.65 | 62 | 25.73 | 19 | 15.57 | .125 |
|  |  | Bachelor | 131 | 33.33 | 80 | 22.1 | 41 | 33.61 |  |
|  |  | Master | 142 | 36.13 | 81 | 22.38 | 50 | 40.98 |  |
|  |  | PhD or higher | 31 | 7.89 | 17 | 4.7 | 12 | 9.84 |  |
|  | Employment |  |  |  |  |  |  |  |  |
|  |  | Employed | 335 | 85.03 | 208 | 86.31 | 102 | 83.61 | .602 |
|  |  | Unemployed or receiving social welfare | 22 | 5.58 | 10 | 4.15 | 4 | 3.28 |  |
|  |  | Retired | 20 | 5.08 | 13 | 5.39 | 7 | 5.74 |  |
|  |  | Student | 17 | 4.31 | 10 | 4.15 | 9 | 7.38 |  |
|  | Migration status |  |  |  |  |  |  |  |  |
|  |  | No migration status | 325 | 82.91 | 200 | 82.99 | 101 | 82.79 | .426 |
|  |  | First generation migrant | 51 | 13.01 | 29 | 12.03 | 18 | 14.75 |  |
|  |  | Second generation migrant | 16 | 4.08 | 12 | 4.98 | 3 | 2.46 |  |
|  | Residence | |  |  |  |  |  |  |  |
|  |  | Rest of the country | 154 | 39.10 | 95 | 39.42 | 49 | 40.16 | .891 |
|  |  | Randstad (main urban area) | 240 | 60.90 | 146 | 60.58 | 73 | 59.84 |  |
| Behavioural determinants | Number of sex partners in the previous 6 months | |  |  |  |  |  |  |  |
|  |  | None | 8 | 2.03 | 1 | 0.41 | 6 | 4.92 | <.001 |
|  |  | 1 | 46 | 11.68 | 13 | 5.39 | 28 | 22.95 |  |
|  |  | 2 to 6 | 82 | 20.81 | 66 | 27.39 | 10 | 8.2 |  |
|  |  | 7 to 15 | 159 | 40.36 | 91 | 37.76 | 59 | 48.36 |  |
|  |  | More than 15 | 99 | 25.13 | 70 | 29.05 | 19 | 15.57 |  |
|  | HIV status |  |  |  |  |  |  |  |  |
|  |  | HIV negative | 363 | 92.13 | 241 | 100,00 | 122 | 100,00 | NA |
|  |  | HIV positive | 22 | 5.58 | 0 | 0,00 | 0 | 0,00 |  |
|  |  | HIV status unknown or not disclosed | 9 | 2.28 | 0 | 0,00 | 0 | 0,00 |  |
|  | PrEP use status |  |  |  |  |  |  |  |  |
|  |  | Current PrEP users | 241 | 66.39 | NA | NA | NA | NA | NA |
|  |  | PrEP naïve or PrEP discontinued | 122 | 30.96 | NA | NA | NA | NA |  |
|  | Ever used any type of substances in the previous 6 months | |  |  |  |  |  |  |  |
|  |  | Never | 349 | 11.42 | 221 | 91.7 | 102 | 83.61 | .020 |
|  |  | Ever | 45 | 11.42 | 20 | 8.3 | 20 | 16.39 |  |
|  | Recreational drugs use in the previous 6 months | |  |  |  | 0,00 |  | 0,00 |  |
|  |  | Never | 250 | 63.45 | 142 | 58.92 | 88 | 72.13 | .014 |
|  |  | Ever | 144 | 36.55 | 99 | 41.08 | 34 | 27.87 |  |
|  | Chemsex in the previous 6 months |  |  |  |  |  |  |  |  |
|  |  | Never | 293 | 74.37 | 152 | 63.07 | 115 | 94.26 | <.001 |
|  |  | Ever | 101 | 25.63 | 89 | 36.93 | 7 | 5.74 |  |
|  | Poppers in the previous 6 months |  |  |  |  |  |  |  |  |
|  |  | Never | 183 | 46.45 | 88 | 36.51 | 82 | 67.21 | <.001 |
|  |  | Ever | 211 | 53.55 | 153 | 63.49 | 40 | 32.79 |  |
|  | Erectile dysfunction medication use in the previous 6 months | |  |  |  |  |  |  |  |
|  |  | Never | 228 | 57.87 | 110 | 45.64 | 99 | 81.15 | <.001 |
|  |  | Ever | 166 | 42.13 | 131 | 54.36 | 23 | 18.85 |  |
|  | Alcohol in the previous 6 months |  |  |  |  |  |  |  |  |
|  |  | Never | 93 | 23.6 | 57 | 23.65 | 29 | 23.77 | .980 |
|  |  | Ever | 301 | 76.4 | 184 | 76.35 | 93 | 76.23 |  |
|  | Visit a gay sauna in the previous 6 months |  |  |  |  |  |  |  |  |
|  |  | Never | 251 | 63.71 | 142 | 58.92 | 89 | 72.95 | .009 |
|  |  | Ever | 143 | 36.29 | 99 | 41.08 | 33 | 27.05 |  |
|  | Visit a darkroom in the previous 6 months |  |  |  |  |  |  |  |  |
|  |  | Never | 238 | 60.41 | 127 | 52.7 | 90 | 73.77 | <.001 |
|  |  | Ever | 156 | 39.59 | 114 | 47.3 | 32 | 26.23 |  |
|  | Visit a circuit party in the previous 6 months | |  |  |  |  |  |  |  |
|  |  | Never | 283 | 71.83 | 162 | 67.22 | 96 | 78.69 | .023 |
|  |  | Ever | 111 | 28.17 | 79 | 32.78 | 26 | 21.31 |  |
|  | Visit a pride event in the previous 6 months |  |  |  |  |  |  |  |  |
|  |  | Never | 203 | 51.52 | 119 | 49.38 | 63 | 51.64 | .684 |
|  |  | Ever | 191 | 48.48 | 122 | 50.62 | 59 | 48.36 |  |
|  | Visit a gay dance club in the previous 6 months | |  |  |  |  |  |  |  |
|  |  | Never | 137 | 34.77 | 74 | 30.71 | 48 | 39.34 | .100 |
|  |  | Ever | 257 | 65.23 | 167 | 69.29 | 74 | 60.66 |  |
|  | Attend private sex parties in the previous 6 months | |  |  |  |  |  |  |  |
|  |  | Never | 277 | 70.3 | 145 | 60.17 | 110 | 90.16 | <.001 |
|  |  | Ever | 117 | 29.7 | 96 | 39.83 | 12 | 9.84 |  |
|  | Visit fetish events/fairs in the previous 6 months | |  |  |  |  |  |  |  |
|  |  | Never | 308 | 78.17 | 174 | 72.2 | 106 | 86.89 | .002 |
|  |  | Ever | 86 | 21.83 | 67 | 27.8 | 16 | 13.11 |  |
| Psycho-social determinants | Knowing anybody who has/had monkeypox | |  |  |  |  |  |  |  |
|  |  | No | 326 | 82.74 | 193 | 80.08 | 107 | 87.7 | .070 |
|  |  | Yes | 68 | 17.26 | 48 | 19.92 | 122 | 100,00 |  |
|  | Cocnern about being infected by monkeypox* | | 4 | [2-4] | 4 | [2-4] | 4 | [2-4] | 0.043 |
|  | Perceived risk of being infected by monkeypox* | | 3 | [2-4] | 3 | [2-4] | 3 | [2-4] | 0.026 |
|  | Perceived problematic consequences of monkeypox* | | 4 | [3-4] | 4 | [3-4] | 4 | [3-5] | 0.206 |

Notes: ^1^ I use substances recreationally (for example THC, MDMA, ecstasy, etc). ^2^ I use substances in the context of sex (for example crystal meth/tina, GHB, ketamine etc.). ^3^ I use erectile dysfunction medication (for example Viagra, Kamagra). * indicates variable with a 1−5 Likert scale, with 1 = extremely unlikely and 5 = extremely likely), results were reported in median [interquartile range]. NA = not applicable.

**Table S2. Sensitivity analysis - determinants of Monkeypox vaccination intention and self-isolation intention**

| **Variables** | | | | **Vaccination intention**  **(Somehow and extremely likely vs. rest of scale)** | | | | | | **Self-isolation intention**  **(Somehow and extremely likely vs. rest of scale)** | | | | | | | | |
| --- | --- | --- | --- | --- | --- | --- | --- | --- | --- | --- | --- | --- | --- | --- | --- | --- | --- | --- |
|  |  |  |  | **Univariable model** | | | **Multivariable model** | | | **Univariable model** | | | | | **Multivariable model** | | | |
|  |  |  |  | **OR** | **95%CI** | **p-value** | **aOR** | **95%CI** | **p-value** | **OR** | | **95%CI** | | **p-value** | **aOR** | | **95%CI** | **p-value** |
| **Socio-demographic determinants** | Age | | |  |  |  |  |  |  |  | |  | |  |  | |  |  |
|  |  | | <45 y.o. | ref. | - | - |  |  |  | ref. | | - | | - |  | |  |  |
|  |  | | >45 y.o. | 1.12 | 0.57;2.21 | 0.743 |  |  |  | 0.84 | | 0.52;1.34 | | 0.461 |  | |  |  |
|  | Relationship | | |  |  |  |  |  |  |  | |  | |  |  | |  |  |
|  |  | | Single | ref. | - | - | ref. | - | - | ref. | | - | | - |  | |  |  |
|  |  | | Single but dating | 2.54 | 0.90;7.11 | 0.077 | 2.66 | 0.87;8.11 | 0.078 | 0.83 | | 0.41;1.69 | | 0.609 |  | |  |  |
|  |  | | Monogamous relationship | 0.72 | 0.26;2.01 | 0.526 | 0.98 | 0.30;3.13 | 0.998 | 0.87 | | 0.34;2.23 | | 0.779 |  | |  |  |
|  |  | | Open/Polyamorous relationship | 2.64 | 1.13;6.17 | 0.025 | 2.42 | 0.96;6.06 | 0.047 | 0.90 | | 0.49;1.65 | | 0.722 |  | |  |  |
|  | Education | | |  |  |  |  |  |  |  | |  | |  |  | |  |  |
|  |  | Lower than Bachelor | | ref. | - | - |  |  |  | | ref. | | - | - | |  |  |  |
|  |  | Bachelor | | 1.40 | 0.59;3.32 | 0.448 |  |  |  | | 0.79 | | 0.42;1.50 | 0.473 | |  |  |  |
|  |  | Master | | 1.40 | 0.60;3.28 | 0.439 |  |  |  | | 1.03 | | 0.56;1.91 | 0.914 | |  |  |  |
|  |  | PhD or higher | | 4.23 | 0.52;34.20 | 0.176 |  |  |  | | 0.89 | | 0.34;2.43 | 0.811 | |  |  |  |
|  | Employment | | |  |  |  |  |  |  |  | |  | |  |  | |  |  |
|  |  | Employed | | ref. | - | - |  |  |  | | ref. | | - | - | | ref. | - | - |
|  |  | Unemployed or receiving social welfare | | 0.82 | 0.18;3.74 | 0.797 |  |  |  | | 1.31 | | 0.45;3.82 | 0.625 | | 1.32 | 0.45;3.88 | 0.613 |
|  |  | Retired | | 464836.00 | 0.00;inf | 0.985 |  |  |  | | 0.15 | | 0.02;1.13 | 0.065 | | 0.15 | 0.02;1.13 | 0.066 |
|  |  | Student | | 0.98 | 0.22;4.43 | 0.987 |  |  |  | | 1.04 | | 0.37;2.96 | 0.933 | | 1.01 | 0.25;2.88 | 0.986 |
|  | Migration status | | |  |  |  |  |  |  |  | |  | |  |  | |  |  |
|  |  | No migration status | | ref. | - | - |  |  |  | | ref. | | - | - | |  |  |  |
|  |  | First generation migrant | | 0.53 | 0.23;1.23 | 0.137 |  |  |  | | 0.56 | | 0.25;1.24 | 0.154 | |  |  |  |
|  |  | Second generation migrant | | 416842.00 | 0.00;inf | 0.988 |  |  |  | | 0.43 | | 0.10;1.93 | 0.271 | |  |  |  |
|  | Residence | | |  |  |  |  |  |  |  | |  | |  |  | |  |  |
|  |  | The rest of the country | | ref. | - | - |  |  |  | | ref. | | - | - | |  |  |  |
|  |  | Randsrad (main urban area) | | 1.54 | 0.78;3.03 | 0.213 |  |  |  | | 0.94 | | 0.58;1.51 | 0.800 | |  |  |  |
| **Behavioural determinants** | Number of sex partners in the previous 6 months | | |  |  |  |  |  |  |  | |  | |  |  | |  |  |
|  |  | None | | ref. | - | - |  |  |  | | ref. | | - | - | |  |  |  |
|  |  | 1 | | 1.58 | 0.27;9.32 | 0.611 |  |  |  | | ***1.05*** | | 0.11;10.10 | 0.966 | |  |  |  |
|  |  | 2 to 6 | | 3.74 | 0.69;20.44 | 0.128 |  |  |  | | 1.76 | | 0.21;14.85 | 0.352 | |  |  |  |
|  |  | 7 to 15 | | 4.38 | 0.74;25.86 | 0.103 |  |  |  | | 2.76 | | 0.32;23.47 | 0.306 | |  |  |  |
|  |  | More than 15 | | 3.57 | 0.60;21.13 | 0.161 |  |  |  | | 3.07 | | 0.36;26.28 | 0.602 | |  |  |  |
|  | HIV status | | |  |  |  |  |  |  |  | |  | |  |  | |  |  |
|  |  | HIV negative | | ref. | - | - |  |  |  | | ref. | | - | - | |  |  |  |
|  |  | HIV positive | | 4254481.23 | 0.00-inf | 0.826 |  |  |  | | 1.24 | | 0.47;3.28 | 0.657 | |  |  |  |
|  |  | HIV status unknown or not disclosed | | 0.13 | 0.03;1.27 | 0.102 |  |  |  | | 0.95 | | 0.19;4.65 | 0.949 | |  |  |  |
|  | PrEP use status | | |  |  |  |  |  |  |  | |  | |  |  | |  |  |
|  |  | Current PrEP users | | ref. | - | - |  |  |  | | ref. | | - | - | |  |  |  |
|  |  | PrEP naïve or PrEP discontinued | | 0.58 | 0.28;1.19 | 0.134 |  |  |  | | 0.74 | | 0.43;1.36 | 0.266 | |  |  |  |
|  | Ever used any type of substances | | |  |  |  |  |  |  |  | |  | |  |  | |  |  |
|  | in the previous 6 months | | |  |  |  |  |  |  |  |  |  |  |  |  |  |  |  |
|  |  | Never | | ref. | - | - |  |  |  | | ref. | | - | - | |  |  |  |
|  |  | Ever | | 1.07 | 0.36;3.18 | 0.902 |  |  |  | | 1.39 | | 0.74;2.58 | 0.302 | |  |  |  |
|  | Recreational drugs use in the previous 6 months ^1^ | | |  |  |  |  |  |  |  | |  | |  |  | |  |  |
|  |  | Never | | ref. | - | - |  |  |  | | ref. | | - | - | |  |  |  |
|  |  | Ever | | *1.62* | 0.76;3.46 | 0.210 |  |  |  | | *0.57* | | 0.25;1.32 | 0.194 | |  |  |  |
|  | Chemsex in the previous 6 months ^2^ | | |  |  |  |  |  |  |  | |  | |  |  | |  |  |
|  |  | Never | | ref. | - | - |  |  |  | | ref. | | - | - | |  |  |  |
|  |  | Ever | | 0.69 | 0.33;1.43 | 0.322 |  |  |  | | 0.85 | | 0.52;1.39 | 0.517 | |  |  |  |
|  | Poppers in the previous 6 months | | |  |  |  |  |  |  |  | |  | |  |  | |  |  |
|  |  | Never | | ref. | - | - | ref. | - | - | | ref. | | - | - | |  |  |  |
|  |  | Ever | | 2.02 | 1.01;4.05 | 0.047 | 1.44 | 0.67;3.12 | 0.355 | | 0.93 | | 0.58;1.48 | 0.762 | |  |  |  |
|  | Erectile dysfunction medication use | | |  |  |  |  |  |  |  | |  | |  |  | |  |  |
|  | in the previous 6 months ^3^ | | |  |  |  |  |  |  |  |  |  |  |  |  |  |  |  |
|  |  | Never | | ref. | - | - |  |  |  | | ref. | | - | - | |  |  |  |
|  |  | Ever | | 0.96 | 0.48;1.88 | 0.886 |  |  |  | | 0.96 | | 0.60;1.54 | 0.854 | |  |  |  |
|  | Alcohol in the previous 6 months | | |  |  |  |  |  |  |  | |  | |  |  | |  |  |
|  |  | Never | | ref. | - | - |  |  |  | | ref. | | - | - | |  |  |  |
|  |  | Ever | | 0.88 | 0.39;2.00 | 0.766 |  |  |  | | 1.63 | | 0.89;2.95 | 0.111 | |  |  |  |
|  | Visit a gay sauna in the previous 6 months | | |  |  |  |  |  |  |  | |  | |  |  | |  |  |
|  |  | Never | | ref. | - | - |  |  |  | | ref. | | - | - | |  |  |  |
|  |  | Ever | | 1.06 | 0.52;2.15 | 0.878 |  |  |  | | 1,01 | | 0.66;152 | 0.965 | |  |  |  |
|  | Visit a darkroom in the previous 6 months | | |  |  |  |  |  |  |  | |  | |  |  | |  |  |
|  |  | Never | | ref. | v | - |  |  |  | | ref. | | - | - | |  |  |  |
|  |  | Ever | | 0.85 | 0.43;1.68 | 0.634 |  |  |  | | 0.81 | | 0.50;1.32 | 0.254 | |  |  |  |
|  | Visit a circuit party in the previous 6 months | | |  |  |  |  |  |  |  | |  | |  |  | |  |  |
|  |  | Never | | ref. | - | - |  |  |  | | ref. | | - | - | |  |  |  |
|  |  | Ever | | 1.07 | 0.50;2.28 | 0.871 |  |  |  | | 0.81 | | 0.48;1.38 | 0.440 | |  |  |  |
|  | Visit a pride event in the previous 6 months | | |  |  |  |  |  |  |  | |  | |  |  | |  |  |
|  |  | Never | | ref. | - | - |  |  |  | | ref. | | - | - | |  |  |  |
|  |  | Ever | | 1.12 | 0.57;2.21 | 0.746 |  |  |  | | 1.14 | | 0.72;1.83 | 0.567 | |  |  |  |
|  | Visit a gay dance club in the previous 6 months | | |  |  |  |  |  |  |  | |  | |  |  | |  |  |
|  |  | Never | | ref. | - | - |  |  |  | | ref. | | - | - | |  |  |  |
|  |  | Ever | | 0.89 | 0.43;1.83 | 0.754 |  |  |  | | 1.48 | | 0.89;2.46 | 0.136 | |  |  |  |
|  | Attend private sex parties in the previous 6 months | | |  |  |  |  |  |  |  | |  | |  |  | |  |  |
|  |  | Never | | ref. | - | - |  |  |  | | ref. | | - | - | |  |  |  |
|  |  | Ever | | 1.16 | 0.54;2.47 | 0.709 |  |  |  | | 1.12 | | 0.68;1.85 | 0.662 | |  |  |  |
|  | Visit fetish events/fairs in the previous 6 months | | |  |  |  |  |  |  |  | |  | |  |  | |  |  |
|  |  | Never | | ref. | - | - |  |  |  | | ref. | | - | - | |  |  |  |
|  |  | Ever | | 0.86 | 0.39;1.89 | 0.700 |  |  |  | | 1.37 | | 0.79;2.35 | 0.260 | |  |  |  |
| **Psycho-social determinants** | Knowing anybody who has/had monkeypox | | |  |  |  |  |  |  |  | |  | |  |  | |  |  |
|  |  | No | | ref. | - | - | ref. | - | - | | ref. | | - | - | |  |  |  |
|  |  | Yes | | 3.97 | 0.93;16.91 | 0.062 | 2.52 | 0.53;11.90 | 0.243 | | 1.12 | | 0.61;2.05 | 0.724 | |  |  |  |
|  | Concern about being infected by monkeypox* | | | 2.32 | 1.68;3.20 | <0.001 | 1.95 | 1.32;2.88 | <0.001 | 0.87 | | 0.72;1.05 | | 0.151 |  | |  |  |
|  | Perceived risk of being infected by monkeypox* | | | 2.21 | 1.55;3.16 | <0.001 | 1.29 | 0.82;2.01 | 0.275 | 0.96 | | 0.77;1.19 | | 0.692 |  | |  |  |
|  | Perceived problematic consequences of monkeypox* | | | 1.37 | 0.99;1.87 | 0.0532 | 1.26 | 0.89;1.78 | 0.192 | 0.80 | | 0.64;1.01 | | 0.062 | 0.80 | | 0.64;1.01 | 0.063 |

Notes: ^1^ I use substances recreationally (for example THC, MDMA, ecstasy, etc). ^2^ I use substances in the context of sex (for example crystal meth/tina, GHB, ketamine etc.). ^3^ I use erectile dysfunction medication (for example Viagra, Kamagra). * indicates variable with a 1−5 Likert scale, with 1 = extremely unlikely and 5 = extremely likely)
